# The effect of the minimum price for unit of alcohol in Scotland on alcohol-related ambulance callouts: a controlled interrupted time series analysis

**DOI:** 10.1101/2022.12.18.22283513

**Authors:** Francesco Manca, Jim Lewsey, Daniel Mackay, Colin Angus, David Fitzpatrick, Niamh Fitzgerald

## Abstract

**Background and aim:** On the 1st of May 2018, to reduce alcohol-related harm, Scotland introduced a minimum unit price (MUP) of £0.50 for alcohol. It has been shown that the policy has reduced per capita alcohol consumption. The aim of this study is to assess the association between the implementation of MUP and changes in the overall volume of alcohol-related ambulance callouts. We then assessed effects on time of the day when alcohol callouts are more numerous and in specific subpopulations (e.g., different socioeconomic groups) to assess potential impacts on health inequality.

**Methods:** A controlled interrupted time series design (ITS) was used to examine variations in the daily volume of alcohol-related callouts (overall and night-time only). The control series was the number of ambulance callouts to under 13 years old. We performed uncontrolled ITS on both the intervention and control group as well as a controlled ITS built on the difference between the two series.

**Results:** No significant decrease in the volume of callouts was found in both uncontrolled series (step change: 0.062, 95%CI: -0.012,0.0135 p=0.091; slope change: -0.001, 95%CI: -0.001, 0.000 p=0.139) and controlled (step change: -0.01, 95%CI: -0.317,0.298 p=0.951; slope change: -0.003, 95%CI: -0.008, 0.002 p=0.257). Similarly, no significant changes were found for the night-time series or for any population subgroups.

**Conclusions:** There was no apparent association between the introduction of MUP and the volume of alcohol-related ambulance callouts. One possible reason for this finding may be the relatively low level of MUP, and its consequent limited effect on alcohol sales.

## Introduction

In 2018, it was estimated that alcohol was responsible for 5.1% of the global burden of disease and injury[1], causing both acute and long-term conditions. Alcohol-related harm, which also includes injuries, violence and other accidents related to binge drinking, also places a significant strain on healthcare and emergency services. Alcohol consumption in Western Europe is among the highest in the world [2], and in the UK 24% of adults regularly exceed drink Chief Medical Officer’s low-risk guidelines[3, 4] and 27% report binge drinking during the heaviest weekly drinking days[5]. In Scotland, there are substantially higher levels of alcohol consumption and alcohol related harm than the rest of the UK [6, 7]. In 2019, 16.2% of ambulance callouts were found to be alcohol-related, with a higher percentage at weekends [8].

In 2018, to reduce consumption and alcohol-related harm in the population, Scotland implemented a minimum unit pricing policy (MUP) for alcohol of £0.50, meaning that one UK unit of alcohol (10ml or 8g of ethanol) cannot be sold below this threshold. As MUP affects only the cheapest alcohol, it was anticipated that the policy would primarily target heavier drinkers [9] As alcohol-related harm is more severe in lower socioeconomic groups, the policy was also expected to have a greater impact on these populations with a consequent decrease in the significant health inequalities that exist in Scotland. After one year of the policy, off-trade alcohol sales were observed to have fallen by 3.5% [10], an effect that was largely sustained at three-year follow-up [11]. As overall consumption decreased, it is reasonable to expect that also some alcohol-related harms would also reduce. Studies to date have found inconclusive effects of MUP on several measures of alcohol harm. For example, MUP was associated with no change in alcohol-related crime [12] or alcohol-related attendances at emergency department[13].

However, many aspects of alcohol-related harm are typically underreported[14, 15], making data from front line emergency services on the role of alcohol in service usage, important for understanding the impact of policies like MUP on harms. The Scottish Ambulance Service is available free at the point of delivery to the whole population of Scotland (∼5.5 million people) as part of the National Health Service. It is often the first and only service in contact with some patients who are treated in the community and who may therefore be missing from A&E and admissions data. As such ambulance callouts may be a ‘more sensitive’ thermometer of the effect of a public health policy than hospital data, especially reflecting the impact of the policy on acute alcohol-related harm. The only other international study analysing the effect of similar MUP policy on ambulance callouts was an interrupted time series study from Australia’s Northern Territory[16]. It reported a significant negative step change but not a significant slope change in the rate of ambulance attendances post-MUP in the overall region, and significant negative slope changes but not step changes in specific local areas. To our knowledge, no published peer review study to date has examined the impact of MUP or any other increase in alcohol prices on ambulance callouts.

This study aimed to identify whether the introduction of MUP in Scotland was associated with changes in the overall volume of alcohol-related ambulance callouts, and whether there were variations across time of the day, the sex, age of the patient, or level of socioeconomic deprivation of the callout location. The legislation that introduced MUP contains a ‘sunset clause’ meaning that it will end after five years of its implementation unless the Scottish Parliament votes for it to continue. This decision will be informed by a large body of evaluation evidence, to which this study will add a new angle.

## Methods

### Study design

We used a controlled, interrupted time series design to evaluate the medium-term impact of the introduction of MUP on alcohol-related ambulance callouts. We used ambulance callouts to patients under 13 years of age, for any cause, as a characteristic-based control as this outcome was not expected to be impacted by the policy [17].

### Dataset

Our data provider was the Scottish Ambulance Service, which supplied a dataset containing selected anonymised fields of all electronic patient clinical records (ePCRs) of ambulance callouts in Scotland from 1^st^ May 2015 to 31^st^ October 2021, covering 3 years prior to and 2.5 years after MUP implementation. Every callout contained information on patients’ age and gender, as well as deprivation decile of the callout location, assessed using the Scottish socioeconomic index of multiple deprivation (SIMD), a composite measure of small area deprivation[18]. The dataset also included two markers of whether or not the call was alcohol-related. Firstly, at the time of completing the records clinicians could select an on/off field to indicate whether alcohol was an ‘additional factor’ in the callout. Secondly, all ePCRs include a free text field into which the ambulance clinician could enter additional notes. The dataset included a yes/no marker generated from an algorithm embedded in the SAS system, which analyses this free text for alcohol-related words and phrases to determine if the callout was alcohol-related. Each individual record was deemed to be alcohol-related for this study if either of these markers were positive for alcohol involvement. This indicator combining the yes/no field with the algorithm field, was developed and validated previously and found to have a sensitivity of 94%, compared to 38% for the paramedic selected on/off field alone. A detailed description of the algorithm development and performance is elsewhere [8].

While the full dataset contained records from 1st May 2015 to 31st October 2021, SAS changed their software system for recording callouts at the end of 2017, with implementation phased in across different Scottish regions over several months. The new recording system gave clinicians multiple options by which they could indicate that alcohol (or other substances) was contributing factor in a given callout, as well as changing the positioning and prominence of such fields. This change had little impact on the free text data which feeds into the algorithm, but the new system increased the number of ways to note alcohol as a contributing factor for the callouts within pre-selected fields by ambulance clinicians. As a result, the implementation of the new system created a gradual increase in the volume of callouts identified by the algorithm as alcohol-related, lasting all the implementation period (a couple of months), followed by a more stable level of alcohol-related callouts from December 2017 (only a few months before the MUP introduction). Such variation in data collection systems could generate structural breaks in the time series, and for this reason, whenever data collection is not consistent over time it is recommended to truncate the analysis period for interrupted time series analyses [19]. Therefore, to avoid potential bias in our analysis, we chose to perform the main analysis only on the period when the new system was fully adopted (after 15^th^ December 2017). The analytical dataset was further truncated in March 2020 to avoid additional bias as a result of the COVID-19 pandemic and its impacts. The lockdown period affected not only alcohol-related ambulance callouts but potentially also consumption patterns [20], with likely long-lasting effects. Consequently, for the main analysis the final dataset was from 15th December 2017 to 15th March 2020.

To calculate the overall effect of MUP on the burden of alcohol to the ambulance service, we would ideally analyse the number of individual patients treated by ambulance crews. However, SAS classifies its incidents in terms of callouts and the data does not record the actual number of patients involved in every callout. Therefore, the unit of measurement in this analysis is “callout”. Whenever we found multiple ePCRs entries referring to one callout, we used the average of patient age. Different records for sex and deprivation within a single callout were very rare, when they happened, they were approximated to the most common record and when there were even numbers they were changed to the most deprived decile and male.

As the characteristic-based control was determined by age (under 13 year olds), all callouts having missing age were removed. A small number of callouts in this control group were identified by the algorithm as being alcohol-related (and therefore fell into both the control and intervention categories) counting for 2.9% of the original alcohol-related callouts. We removed these records from the analysis. As these overlapping observations were uniformly distributed pre- and post-MUP introduction, this is unlikely to have any significant impact on our results. Together these exclusions counted for 8.1% and 9.5% of the original alcohol-related and under 13 years old callouts (Table 1).

**Table 1.** Number of alcohol-related and control (under 13 years old) callouts by demographics. 15th December 2017 to 15th March 2020.

|  | Alcohol (%) |  | Under 13 (%) |  |
| --- | --- | --- | --- | --- |
|  | 190177 |  | 58919 |  |
| **Socioeconomic deprivation quintiles** |  |  |  |  |
| 1 (most deprived) | 66,399 | (35.2%) | 17360 | (30.0%) |
| 2 | 47,436 | (25.1%) | 12866 | (22.3%) |
| 3 | 35,108 | (18.6%) | 10814 | (18.7%) |
| 4 | 24,337 | (12.9%) | 9268 | (16.0%) |
| 5 (east deprived) | 15,577 | (8.2%) | 7504 | (13.0%) |
| Missing | 1,320 |  | 1107 |  |
| **Sex** |  |  |  |  |
| Female | 69,788 | (37.9%) | 22,010 | (42.5%) |
| Male | 114,318 | (62.1%) | 29,776 | (57.5%) |
| Missing | 6,071 |  | 7,133 |  |
| **Age** |  |  |  |  |
| 13-25 | 28,966 | (15.7%) | - |  |
| 26-45 | 52,831 | (28.6%) | - |  |
| 46-65 | 59,010 | (32.0%) | - |  |
| >65 | 43,771 | (23.7%) | - |  |
| Missing | 5,599 | (3.0%) | - |  |
| Sample size after removing callouts with missing age | 180355 |  | 58919 |  |
| Sample size after removing under13 classified as alcohol | 174756 |  | 53320 |  |

### Statistical analysis

We fitted a seasonal autoregressive moving average (SARIMA) model, able to account for autocorrelation, seasonality, and underlying temporal trend. Our main model analysed the potential for a linear change both in both level and slope at the point of intervention. We based this on a potential hypothesis of a gradual effect (in case there was one), that was then tested with information criteria (AIC and BIC statistics) of separate models. The step change in the level of callouts was tested for by fitting a dummy variable corresponding to a date being before 1st May 2018 or after it. The change in trend was given by adding into the model an additional linear trend starting after 1st May 2018 [21] (see Equation 1). We used daily units of time. While a daily series may present challenges (e.g., double seasonality, both weekly and yearly), aggregation to weekly, or monthly data points would have significantly reduced the statistical power of the analysis, due to the restricted size of the original dataset following truncation for changes in reporting and the impact of the pandemic. The use of daily data also allowed us to control for potential time-varying confounders such as weather which is a factor likely to have a role in alcohol consumption [22]. As we were assessing the overall weather for Scotland which has sensitive difference within its territory, to have a single national figure attributable to Scotland, we averaged the daily mm of rainfall in different Scotland districts with data from the Scottish Environment Protection Agency (www.sepa.org.uk) was computed. Other variables included in the model were: bank holidays, months, new year’s eve and Old Firm football matches. Old Firm matches are those between Celtic and Rangers, major football clubs based in Glasgow with a strong sporting rivalry. These were included in the model as there is evidence of increases of domestic violence associated with such matches, with alcohol consumption identified as a potential contributing factor [23]. We included this as a covariate as the metro area population of Glasgow is a relevant proportion of the overall Scottish population and, moreover, Celtic/Rangers fans are present across the country. To reduce the skewness of our original data, we log transformed our dependent variable. An advantage of the log transformation is that coefficients in the regression model can be interpreted as percentage variation of the series. The SARIMA models in our study had the following form: SARIMA(p,d,q)(P,D,Q)_m_ The first bracket describes the non-seasonal autoregressive component where the p,d,and q are the autoregressive, differencing and moving average terms, respectively. The second bracket is the seasonal component with m indicating the seasonal frequency. The most appropriate autoregressive specification depends on series and errors autocorrelation and is series specific. However, for illustrative purposes we report here the equation of the model specification of the alcohol-related callouts, which had the form of SARIMA(1 8,0,0)(2,0,1)_7_:

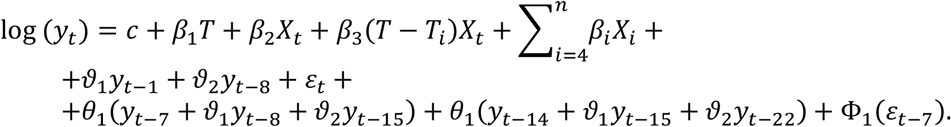

In the first line of the equation, y_t_ is the number of alcohol-related callout at time t, T is the time from the start of the study, X_t_ is the dummy variable representing the step-change, T_i_ represents the time point when MUP started (β_3_ is the change in slope), β_i_ and X_i_ represent exogenous variables (e.g., weather, bank holidays and other), terms in the second line are the non-seasonal autoregressive terms and the random error at time t, terms in the third line are seasonal and moving average components of frequency 7 days.

We performed the same analysis for our characteristic based control outcome. With both control and intervention time series covering the same geographical area (Scotland), they are therefore affected by the same environmental and other unmeasured confounding factors. After testing common pre-trend assumptions, we ran a separate analysis on the difference between control and intervention series which, by incorporating the control into the same model as the intervention analysis, can be interpreted as a difference in difference estimator. Finally, we performed the same analysis for night-time callouts (8pm-6am) as it was the time of the day with the highest concentration of alcohol-related callouts [8].

### Subgroup and sensitivity analysis

Subgroup analysis on different socio-economic deprivation quintiles[24], age groups (13-25, 26-45, 46-65, >65) and sex were performed on the intervention series.

Alternative modelling strategies using panel data and regressions with Newey–West standard errors based on Scottish districts were also employed, using district level covariates (e.g. rainfall levels and old firm matches). However, several areas had low numbers of callouts, including multiple dates with 0 events (with consequent potential floor effects) and challenges in representing correlation and seasonal variations made SARIMA models preferred for baseline analysis.

Falsification tests 6 months and 1 year after the intervention date were performed. Whenever the actual analysis provides statistically significant results, but these are mirrored by significant results in falsification tests, findings can be rejected as causal as influenced by external confounders. In a further sensitivity analysis, to use all the original information of the dataset prior to March 2020, allowing for a longer pre-intervention period, we employed a cubic spline model [25] to mitigate the fluctuation in callouts given by changes in the recording system. Lastly, further sensitivity analyses were run changing the level of approximation of regarding sex and deprivation group.

## Results

**Figure 1.**
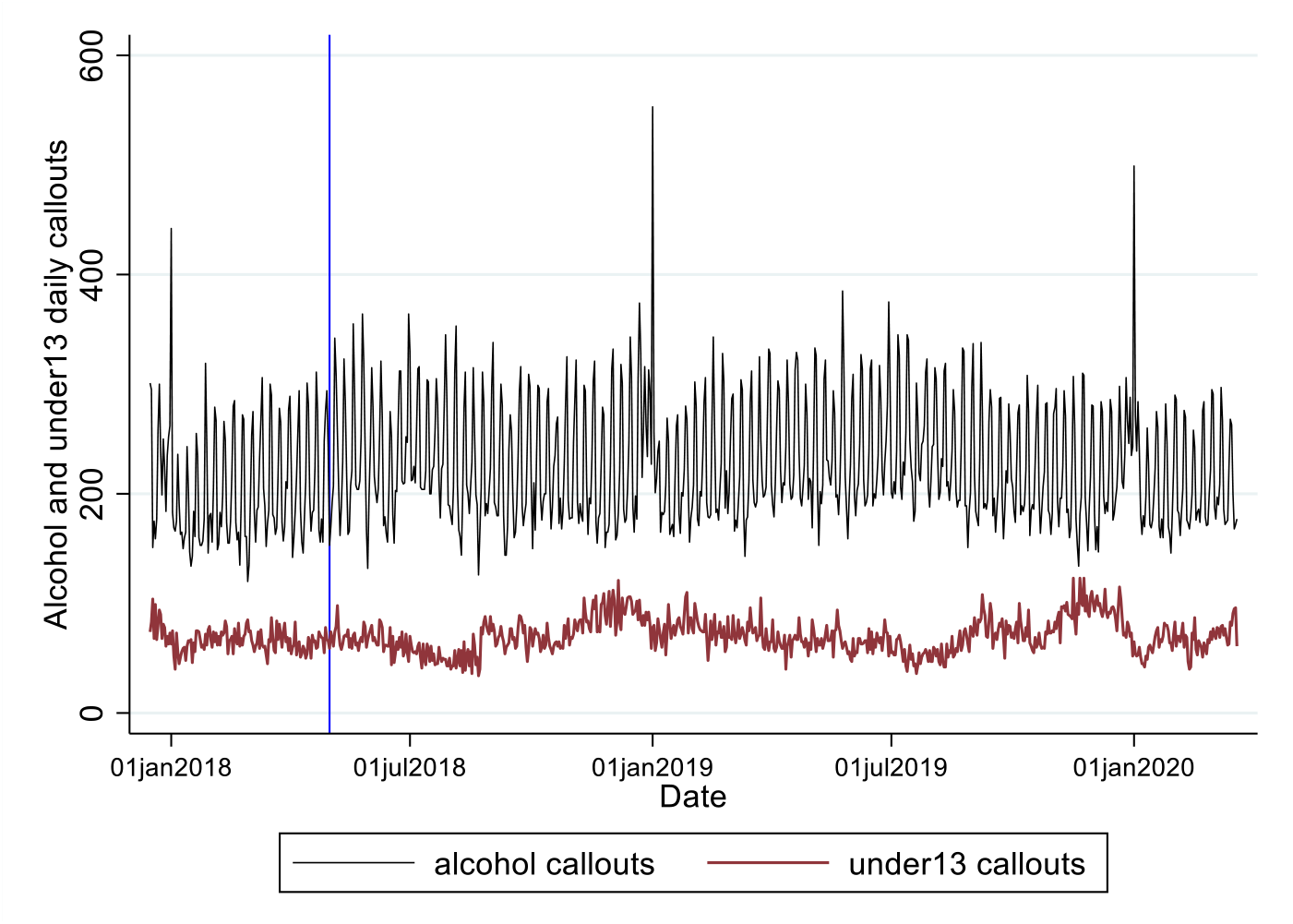
Time series of daily alcohol-related and control outcome (under 13 years old) callouts

Alcohol-related callouts follow a seasonal pattern with peaks at weekends and large peaks on New Year’s Eve (Figure 1). In contrast, callouts for under 13 year olds have little variation over the week, but follow more of a monthly seasonal pattern with gradual growth in November and December and a reduction in the summer months. The overall distribution of sex and socio-economic deprivation was similar in the two groups with males (62% intervention, 57% control) and locations in the most socio-economically deprived fifth of areas (35% vs. 29%) having the highest percentage of callouts (Table 1).

Whilst the mean number of daily alcohol-related callouts before MUP was implemented was 194.3 (SD=51.0) and after implementation was 216.3 (SD=52.4), a relative increase of 11.3%, the inferential analysis, considering seasonal and temporal trends, found that the implementation of MUP was not associated with a significant change in daily alcohol-related callouts (step change -interpretable as an instant change in correspondence to the intervention-: 0.062, 95%CI: -0.012,0.0135 p=0.091; slope change, the daily gradual change after intervention:-0.001, 95%CI: -0.001, 0.000 p=0.139) (Table2). For the control group, the mean number before MUP implementation was 68.4 (SD=10.8) and 72.3 after (SD=16.2), a 5.7% growth, with the inferential analysis also finding no significant step change (step change: -0.006, 95% CI: -0.192, 0.205 p=0.949, slope change: -0.001, 95%CI: -0.003, 0.002 p=0.513).

Similarly, the difference between the intervention and control group did not have significant results (step change: -0.010, 95% CI: -0.317, 0.298 p=0.951, slope change: -0.003, 95%CI: -0.008, 0.002 p=0.257). There were no significant results when the analysis was restricted to alcohol-related callouts at night-time only. In both the main analyses, on total and night-time alcohol-related callouts, the slope change tended to be of the same size, but opposite direction, of the overall trend in the model, indicating a general flat slope after MUP implementation, meaning that the volume of callouts remained stable.

### Subgroup and sensitivity analysis

We found no evidence of a significant decrease in alcohol-related callouts associated with MUP for any of the subgroups examined (different age groups, gender, or callouts to locations with different levels of deprivation). These findings are presented in supplementary material. Where there were statistically significant changes, these happened only for measures at mid ranks of a category (e.g. variations in 3^rd^ and 4^th^ socioeconomic deprived quintiles but not in the least or most deprived groups), we believe these are likely to arise from daily spurious variations. Similarly, significant step changes (in female patients, callout locations in the most deprived quintile, and those aged 46-65) on the day of the implementation of MUP are more likely to represent spurious noise in the data rather than attributable to an instant effect on the first day of MUP. Analogous to the main analysis, in most of the subgroups changes in slope after MUP were often of the same extent of the underlying trend, determining a stable post intervention volume of callouts.

Falsification tests for total alcohol-related callouts produced significant change in slope and trend when the intervention point was set at 6 months after the introduction of MUP, and a significant change in slope, trend and overall underlying trend when set at 12 months after (Table 2). In contrast, falsification tests for the control and the difference between series were not significant. Regarding the night-time analysis, both 6 and 12 month falsification tests in the uncontrolled intervention series had significant results for overall trend and slope change. In the difference between series only the falsification test postponing the intervention by 6 months had significant coefficients for step change (−0.1429 95%CI: - 0.2635 -0.0223 p=0.020), slope change (−0.001 95%CI: -0.001 -0.000 p=0.009) and overall trend (0.001 95%CI: 0.00016 0.0012 p=0.010) with an expected gradual effect at the end of follow up equal to a 35% decrease. This could be interpreted as a lagged gradual decrease in the volume of night-time alcohol-related callouts after 6 months following the introduction of the policy, however given the different sign on the change in slope coefficient in the 12 month falsification test, a more likely hypothesis may be that this is a spurious effect.

**Table 2.**
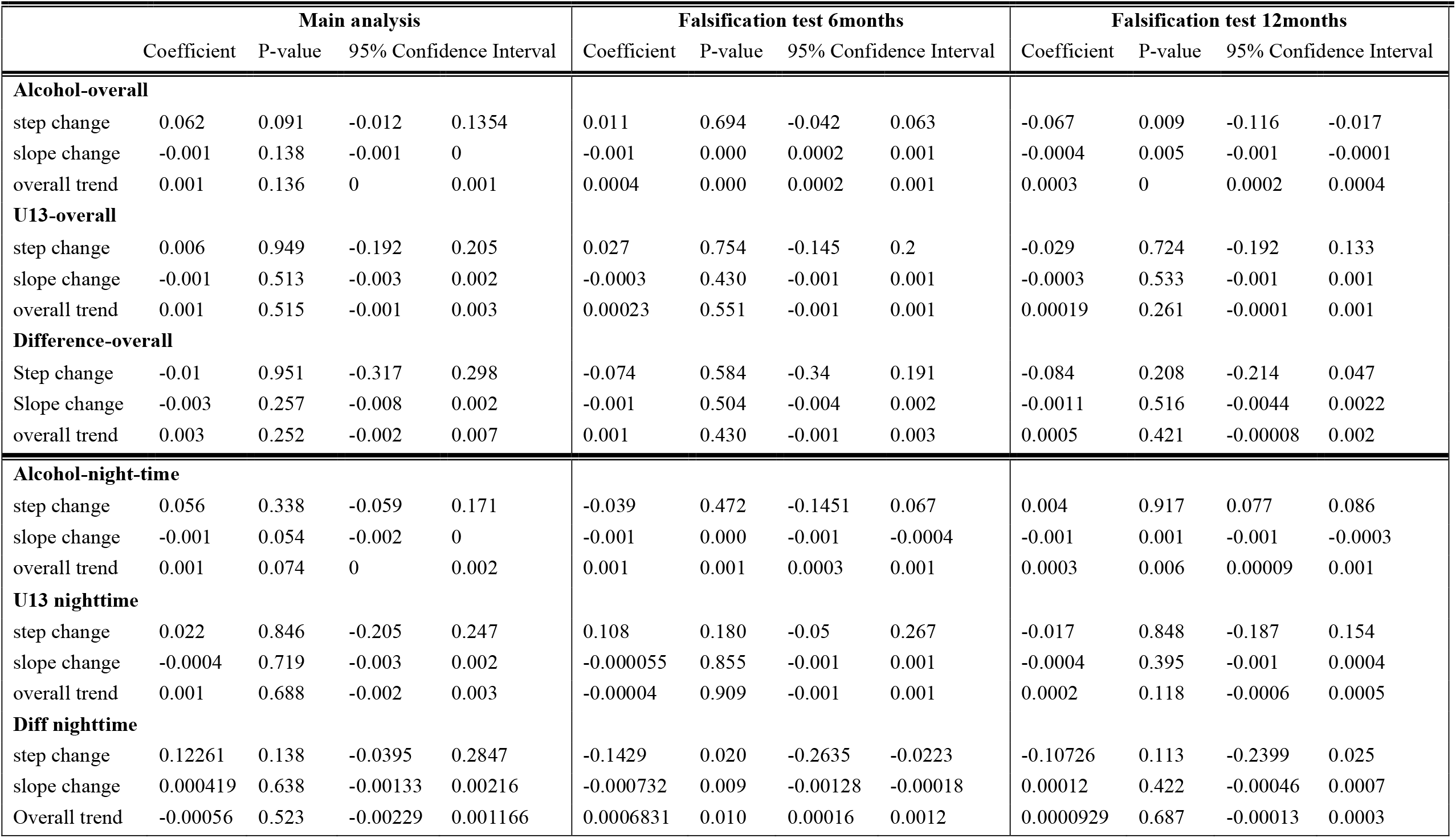
Regression outputs of logarithm of alcohol related callouts (, under 13 years of age callouts and difference between the two series.

The sensitivity analysis using cubic spline starting from May 2015 showed no significant results either and considering also the period when the recording system changed presented a lower goodness of fit. Alternative models such as Newey–West with heteroskedastic and autocorrelated errors did not show significant results in both main analysis and in the difference between the series (supplementary material).

## Discussion

We did not find associations between MUP implementation and a decrease in alcohol-related callouts. There was also no evidence of significant decrease in subpopulations and, in particular, across different socioeconomic groups (relevant sub-analyses to assess the MUP effects on health inequality).

There could have been several reasons why we did not detect a significant association between MUP introduction and changes in the volume of alcohol-related ambulance callouts. We discuss four here. Firstly, the actual level of MUP may have been too low to have an effect on alcohol-related ambulance callouts large enough to be detected, as it was introduced at the same level as when it was first proposed in 2012 without being indexed to inflation, so policy expectations were linked to the real value of £0.50 in 2012. Secondly, many alcohol-related ambulance callouts are generated by alcohol consumption during weekends and night-times [8] much of which takes place in licensed premises (bars, clubs), and MUP does not affect the price of alcohol sold in such premises. However, many people also increase their alcohol consumption at home on weekend nights, and patrons of the night-time economy may also often drink off-trade alcohol in domestic settings prior to going out, so one might have expected to see some effect on ambulance callouts if such consumption decreased. We did find a potential lagged effect of MUP in a decrease in alcohol-related callouts at nighttime after 6 months, but we believe this was most likely a spurious finding (see below).

Thirdly, the kinds of drinking or people that end up generating alcohol-related ambulance call-outs may not be as price-elastic as other alcohol consumption. Whilst overall alcohol consumption fell due to MUP [10, 11], at this level of MUP, perhaps people retained their consumption on ‘big nights’ but instead cut consumption across the week, or perhaps consumption fell predominantly in those groups who are least likely to need an ambulance due to their drinking. Further, many alcohol-related callouts are to individuals with chronic alcohol problems, on whom MUP at the current rate has been found to have limited effect[26]. Understanding what percentage of alcohol-related ambulance callouts are linked to off-trade versus on-trade consumption, and to single occasion versus dependent drinking would help to unpick what happened. Such data are not available in Scotland, though there have been initiatives in emergency departments to identify where people were drinking prior to an alcohol related visit[27]. Finally, the overall reduction in consumption due to MUP may have been generated by very small reductions by a large population of drinkers. Whilst this would have long-term benefits in terms of reduced alcohol-related disease (e.g. cancers), which would be expected to reduce pressure on the health service, it would have no effect on our outcome measure in this study. Ambulance call-outs to chronic diseases partly attributable to alcohol are improbable to be detected as alcohol-related by our algorithm, and would be unlikely to be change in the timeframe of this study.

The fundamental relationship by which less alcohol is purchased when the price rises is not called into question by this study, and is supported by other findings[10, 11], however, it gives cause to consider at what level the minimum price should be set, if the aim is to quickly reduce health service usage and demand. Whilst ambulance callouts are generally responding to acute needs, many are also to patients with chronic alcohol problems; any effects of MUP were not large enough to be detectable in the timeframe or power of our study.

A significant increasing underlying trends in alcohol callouts in subgroups may have appeared because our analysis started in December and included only a few months (not an entire year) before MUP implementation. Therefore, a short pre intervention period could limit the analysis trend which shows higher variability (standard deviations) after MUP implementation for both series. This could explain why the effect of the underlying trend is always offset by the slope change, which describes a flat curve after MUP. We found several step changes in our subgroup analyses as well as in some falsification tests, however, it is worth noticing that we used daily data which are subjected to more noise due to the high level of granularity. To avoid false conclusions, we focused more on significant changes in slope (which imply a continuous change over time from the intervention) rather than on step changes. When falsification tests provided significant results in the overall uncontrolled alcohol-related series, we found no evidence of effect in the series of the differences and similar results (but not statistically significant) in the control (under 13) series. The lack of significance may be due to power issues as the number of under 13 callouts was lower than the number of alcohol-related ones. Therefore, we would not force the interpretation of the significance of our falsification tests as a delayed effect of the policy. In contrast, regarding nighttime callouts, we found a significant decrease in falsification tests after 6 months both in the uncontrolled series and in the series of the difference, while no effect was found in the control group. This could be interpreted as a lagged effect of the policy, but the 12 months falsification test found a non-significant increase in such callouts. It may be that any lagged impact was small, but this was not reflected by the 35% expected effect at the 6 month point. Another possibility is that these results were affected by daily noise and/or by the use of a sub-optimal control group (see limitations below). Overall therefore, we do not interpret this finding as evidence of a delayed effect of the policy.

We believe that our study provides a valuable contribution to evaluating the impact of MUP in Scotland by focusing on its impact on ambulance callouts, that is, on an acute and critical frontline emergency service. The use of a reliable measure of alcohol-related ambulance callouts across the whole of Scotland is a strength, however, our study also presents several limitations. Most importantly, the power of the study was limited by the short pre-intervention period due to the change in the recording system that happened in 2017 and the consequent use of daily data we extensively explained earlier. Other limitations can be related to our control group. We found different seasonal patterns between the intervention and control series (alcohol-related callouts followed weekly fluctuations with peaks during weekends reflecting drinking behaviours, while under 13 callouts had a yearly trend that more closely matches influenza seasons (Figure 1)). In addition, certain covariates (e.g., weather) affected the two series differently, suggesting that the two populations were intrinsically different. However, this was the best control available to us, as a location-based controls such as that used in similar analyses on MUP in Scotland[10, 12] would have been difficult. For instance, there are multiple ambulance trusts in the rest of the UK, and none have embedded a reliable algorithm or marker for identifying which callouts are alcohol-related[28]. While not ideal, our characteristic-based control accounted for local variations such as weather, specific bank holidays, features within ambulance service such as changes in ambulance service provision and the NHS funding environment.

## Conclusions

Minimum unit pricing for alcohol, set at a rate of £0.50 per UK unit of alcohol (8g) and implemented in Scotland in May 2018 was not associated with changes in the volume of alcohol-related ambulance callouts. Further no reduction in such callouts was associated with MUP for any subgroups analysed, including different genders, ages or the level of deprivation of the callout location. The relatively low MUP rate, with limited impact on dependent drinkers and on prices in bars/clubs, most likely explains these findings.

## Data Availability

Most of the Data produced in the present study are stored safely at the University of Glasgow. They are not available for disclosure reasons as provided by the Scottish Ambulance Service. Other data produced in the present work are contained in the manuscript.

## Acknowledgements

This study was funded by CSO (Chief Scientist Office – Scotland-). Grant n.: HIPS 18/57

