## Supplementary material for "The effect of the minimum price for unit of alcohol in Scotland on alcohol-related ambulance callouts: a controlled interrupted time series analysis"

|  | Coefficient | | | P-value | | 95% CI | |
| --- | --- | --- | --- | --- | --- | --- | --- |
| *subgroup analysis* | | | | | | | |
| **Sex** | | | | | | | |
| male | |  |  | |  | |  |
| step change | | 0.046 | 0.321 | | -0.045 | | 0.136 |
| slope change | | -0.001 | 0.185 | | -0.002 | | 0.001 |
| overall trend | | 0.001 | 0.204 | | 0 | | 0.002 |
| female | |  |  | |  | |  |
| step change | | 0.108 | 0.012 | | 0.0238 | | 0.192 |
| slope change | | -0.0001 | 0.846 | | -0.001 | | 0.001 |
| overall trend | | 0.0001 | 0.8 | | -0.001 | | 0.001 |
| **Socioeconomic quintiles** | | | | | | | |
| 1 (most deprived) | |  |  | |  | |  |
| step change | | 0.114 | 0.002 | | 0.041 | | 0.185 |
| slope change | | 0.001 | 0.233 | | -0.0003 | | 0.001 |
| overall trend | | -0.0004 | 0.231 | | -0.001 | | 0.0003 |
| 2 | |  |  | |  | |  |
| step change | | 0.052 | 0.247 | | -0.036 | | 0.14 |
| slope change | | -0.001 | 0.236 | | -0.002 | | 0 |
| overall trend | | 0.001 | 0.205 | | 0 | | 0.002 |
| 3 | |  |  | |  | |  |
| step change | | 0.035 | 0.539 | | -0.078 | | 0.148 |
| slope change | | -0.002 | 0.002 | | -0.003 | | -0.001 |
| overall trend | | 0.002 | 0.002 | | 0.001 | | 0.003 |
| 4 | |  |  | |  | |  |
| step change | | 0.119 | 0.169 | | -0.05 | | 0.288 |
| slope change | | 0.0003 | 0.714 | | -0.002 | | 0.002 |
| overall trend | | -0.0003 | 0.760 | | -0.0022 | | 0.0016 |
| 5 (least deprived) | |  |  | |  | |  |
| step change | | 0.007 | 0.938 | | -0.158 | | 0.1711 |
| slope change | | -0.001 | 0.587 | | -0.003 | | 0.002 |
| overall trend | | 0.001 | 0.523 | | -0.001 | | 0.003 |
| **Age group (years)** | | | | | | | |
| 13-25 | |  |  | |  | |  |
| step change | | 0.04 | 0.7 | | -0.165 | | 0.246 |
| slope change | | -0.002 | 0.124 | | -0.005 | | 0.001 |
| overall trend | | 0.002 | 0.148 | | -0.001 | | 0.004 |
| 26-45 | |  |  | |  | |  |
| step change | | 0.075 | 0.149 | | -0.027 | | 0.178 |
| slope change | | -0.0005 | 0.49 | | -0.002 | | 0.001 |
| overall trend | | 0.0004 | 0.547 | | -0.001 | | 0.002 |
| 46-65 | |  |  | |  | |  |
| step change | | 0.146 | 0.018 | | 0.025 | | 0.268 |
| slope change | | -0.00007 | 0.931 | | -0.0017 | | 0.0015 |
| overall trend | | 0.0000299 | 0.969 | | -0.002 | | 0.002 |
| >65 | |  |  | |  | |  |
| step change | | 0.038 | 0.579 | | -0.096 | | 0.172 |
| slope change | | 0.001 | 0.469 | | -0.001 | | 0.002 |
| overall trend | | -0.0004 | 0.611 | | -0.002 | | 0.001 |

| Newey-West heteroscedastic and autocorrelationconsistent (HAC) standard errors (SEs), assuming autocorrelation up to lag 7, results in logaroithm -exclusion of 3 areas (Wick, Shetlands and Harris) with 0 callouts for several days- | | | | |
| --- | --- | --- | --- | --- |
|  | **Main analysis** | | | |
| Coefficient | | P-value | 95% Confidence Interval | |
| **Alcohol related** |  |  |  |  |
| step change | -0.002 | 0.980 | -0.1561 | 0.1521 |
| slope change | -0.001 | 0.308 | -0.0026 | 0.0008 |
| overall trend | 0.001 | 0.302 | -0.0008 | 0.0027 |
| **Difference** |  |  |  |  |
| step change | 0.006 | 0.921 | -0.1166 | 0.1291 |
| slope change | -0.001 | 0.134 | -0.0025 | 0.0003 |
| overall trend | 0.001 | 0.123 | -0.0003 | 0.0025 |
